# Protocol for a prospective accuracy study on an artificial intelligence-based ultrasound system for gestational age estimation among pregnant women in Ghana, Kenya and South Africa

**DOI:** 10.64898/2026.02.12.26346216

**Authors:** Alim Swarray-Deen, Annie RA McDougall, Rosa Chemway, Rachel Craik, Skandarupan Jayaratnam, Naima Joseph, Robert K Mahar, Digsu N Koye, Long Nguyen, Julie A Simpson, George Gwako, Ruth Hadebe, Edmund Tetteh Nartey, Nicole Minckas, A Metin Gülmezoglu, Joshua P Vogel, Ayesha Osman, PEARLS Collaborators

**Affiliations:** Department of Obstetrics and Gynaecology, University of Ghana Medical School, Accra, Ghana; Women’s, Children’s and Adolescents’ Health Program, Burnet Institute, Melbourne, Australia; Department of Obstetrics and Gynaecology, University of Nairobi, Nairobi, Kenya; Boston Medical Centre, Boston, USA; Centre for Epidemiology and Biostatistics, Melbourne School of Population and Global Health, University of Melbourne, Melbourne, Australia; Methods and Implementation Support for Clinical and Health (MISCH) research Hub, University of Melbourne, Melbourne, Australia; Clinical Epidemiology and Biostatistics Unit, Murdoch Children’s Research Institute, Parkville, Victoria, Australia; University of Cape Town, Cape Town, South Africa; Gender and Women’s Health Unit, Nossal Institute for Global Health, School of Population and Global Health, University of Melbourne, Melbourne, Australia; Concept Foundation, Geneva, Switzerland□ and Bangkok, Thailand

**Keywords:** artificial intelligence, aspirin, gestational age estimation, hypertensive disorders of pregnancy, point of care, pre-eclampsia, protocol, prospective cohort, sub-Saharan Africa, ultrasound

## Abstract

**Background:** Risk screening for pre-eclampsia relies on accurate gestational age assessment, but routine access to ultrasound-based gestational dating remains challenging in many low- and middle-income countries (LMICs). As part of the formative work for the “Preventing pre-eclampsia: Evaluating AspiRin Low-dose regimens following risk Screening” (PEARLS) trial, we aim to validate and implement an Artificial Intelligence (AI)-based algorithm for estimation of gestational age, using blind sweeps done with a handheld ultrasound device. This study protocol outlines the accuracy cohort for AI-based gestational age estimation in participating facilities in Ghana, Kenya, and South Africa.

**Methods:** This multi-country prospective cohort study will recruit 969 pregnant women at 13 health facilities across Kenya, Ghana and South Africa. The eligible population are pregnant women presenting for antenatal visit from 11^+0^ to 13^+6^ weeks’ gestation. Eligible women will have a gestational age assessment by a trained sonographer using fetal biometry (reference standard), followed by gestational age estimation conducted by a trained midwife using the AI-based Intelligent Ultrasound ScanNav FetalCheck system (experimental). Both conventional and AI-based gestational age scans will be conducted with the General Electric (GE) VScan^TM^ Air platform. Women will return for a second visit between 14^+0^ and 27^+6^ weeks’ gestation (week of visit is randomly selected) for an assessment with both conventional and AI-based ultrasound. The primary objective is to determine the accuracy and precision of gestational age estimation using an AI ultrasound system in first and second trimesters, as compared to gestational age estimation using crown-rump length (CRL) measurement by conventional ultrasound in first trimester (11^+0^ to 13^+6^ weeks’).

**Discussion:** The study will provide critical evidence on the accuracy of a point-of-care, AI-based gestational age estimation ultrasound algorithm in sub-Saharan African settings. This study will inform the design of the PEARLS trial, as well as provide vital evidence for expanding implementation of ultrasound-based gestational age assessment for women in Africa.

## Background

Pre-eclampsia remains a leading cause of maternal morbidity and mortality globally, causing an estimated 46,000 maternal deaths annually, over 90% of which occur in low- and middle-income countries (LMICs).^1^ Pre-eclampsia is also a major cause of maternal and neonatal morbidity, including maternal kidney and liver dysfunction, stroke, and seizures, and fetal growth restriction, stillbirth and preterm birth.^2^ Preventive measures, such as identifying women at high risk of developing pre-eclampsia and offering them daily aspirin exist, but are poorly implemented. Many of these deaths could be prevented by implementing systematic risk screening of all women, and risk stratification-based care pathways and preventive measures. The World Health Organization (WHO) recommends that all women who are at moderate or high risk of developing pre-eclampsia should be offered low-dose aspirin for prevention, ideally prior to 20 weeks’ gestation.^3^ However, questions remain about the optimal aspirin dose for this indication. WHO’s recommendations currently favour a 75mg aspirin dose, but the panel highlighted uncertainty around aspirin dosing.^3^ While acknowledging that 150mg might be more effective (though confidence intervals are overlapping), they expressed concern about the possible increased risk of postpartum haemorrhage (PPH) at higher aspirin doses. A comparative trial of 150mg vs 75mg aspirin was recommended, to resolve this uncertainty.

The PEARLS Trial (<u>P</u>reventing pre-eclampsia: <u>E</u>valuating <u>A</u>spi<u>R</u>in <u>L</u>ow-dose regimens following risk <u>S</u>creening) addresses this question through an individually randomised, double-blind trial of pregnant women at high risk of pre-eclampsia in Ghana, Kenya, South Africa and India (PACTR202403785563823; CTRI/2025/08/093004). The PEARLS trial will evaluate whether 150mg is more beneficial than 75mg for preventing preterm pre-eclampsia (superiority hypothesis), with no additional safety concerns, measured by a composite outcome of postpartum haemorrhage (PPH)-related management (non-inferiority hypothesis). Before the PEARLS trial can be implemented, we need to determine the best way to identify women at high-risk of pre-eclampsia. In a formative phase, we are validating a ‘restricted variable’ Fetal Medicine Foundation (FMF) tool for predicting preterm pre-eclampsia in pregnant women in Ghana, Kenya and South Africa, followed by a second cohort in India (Figure 1).^4,5^

**Figure 1.**
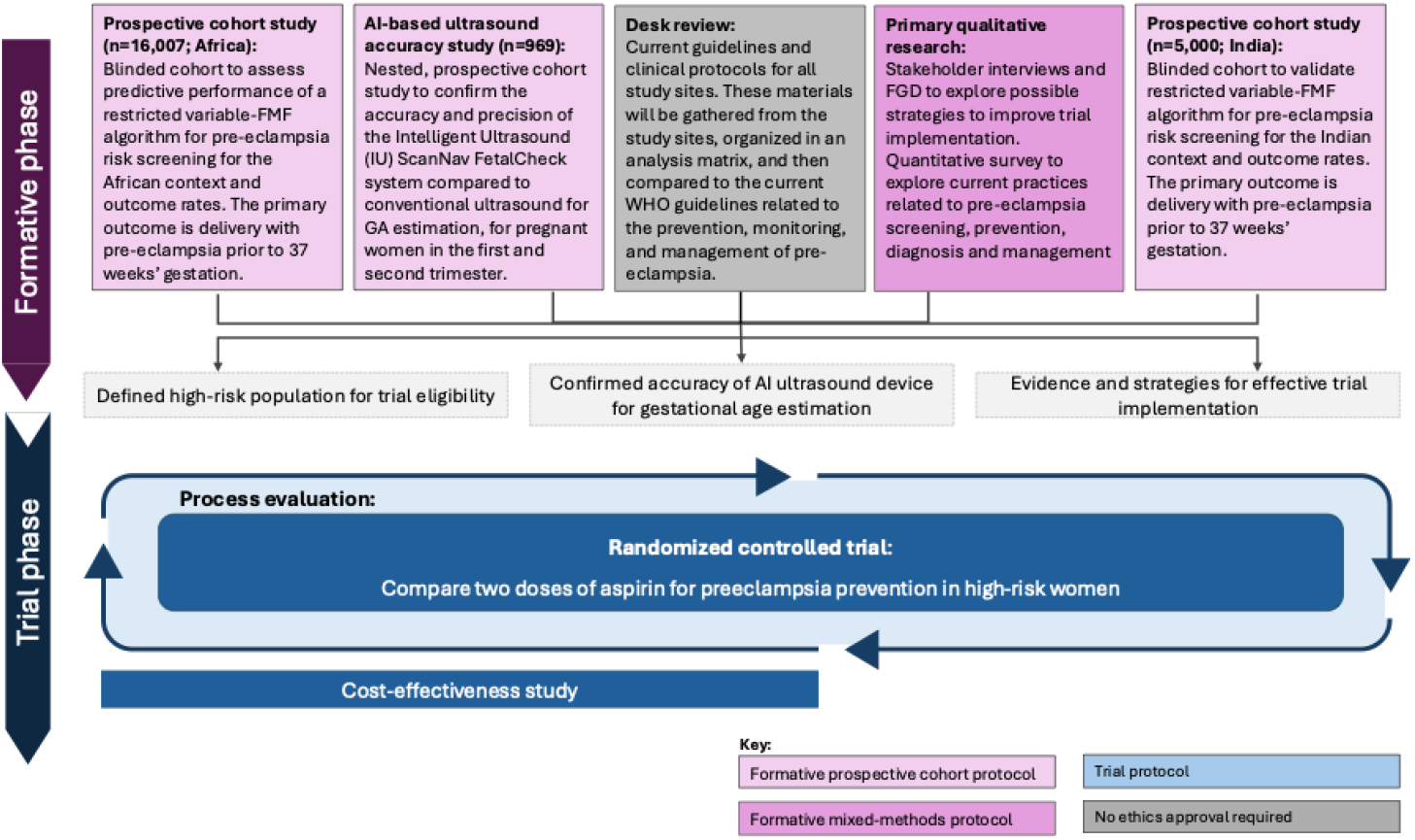
Overview of the PEARLS research activities (reproduced from^5^). The prospective AI-based ultrasound accuracy study is outlined in this protocol. AI = artificial intelligence, FGD = focus group discussions, FMF = Fetal Medicine Foundation, GA = gestational age, IU = Intelligent Ultrasound, WHO = World Health Organization.

The FMF tool uses a Bayesian model combining information on demographic and clinical history-based risk factors, gestational age, as well as mean arterial pressure (MAP), uterine artery Doppler pulsatility index (UTPI), placental growth factor (PLGF) levels and/or pregnancy-associated placental protein A (PAPP-A) levels.^6,7^ In European settings using a risk threshold of ≥1 in 100, the FMF tool has a detection rate (sensitivity) of 80.7% for preterm pre-eclampsia (<37 weeks’ gestational age), with a false positive rate of 14.1%.^7^ Even in the absence of the UTPI and blood-based biomarkers, the detection rate is 66.7%, with a false positive rate of 17.9%.^7^ The FMF tool has been validated in Western European and Asian populations,^8^ and has been assessed in a single tertiary facility in India.^9^ However, the FMF tool has not been validated or implemented at scale in African or South Asian countries. Certain variables within the FMF tool are not routinely available in antenatal care in many LMICs, particularly serum biomarkers, uterine artery Doppler and reliable gestational age estimation using ultrasound. Accurate dating is critical to the provision of risk screening for pre-eclampsia using the FMF-based approach, and in turn implementing a strategy of pre-eclampsia prevention using low-dose aspirin.

### Ultrasound for gestational age assessment: essential but out of reach in many LMICs

WHO recommends that all pregnant women should be offered an antenatal ultrasound before 24 weeks gestation, stating that ultrasound-based estimation of gestational age, and detection of fetal anomalies and multiple pregnancies, improve women’s antenatal care and timing of birth, and may improve women’s pregnancy experience and pregnancy outcome.^10^ It also helps ensure appropriate use of gestational age-specific interventions, such as initiating low-dose aspirin for women at risk of pre-eclampsia prior to 20 weeks’ gestation.^3^ Implementation of antenatal ultrasound for all pregnant women relies on availability of ultrasound devices facilities in which to conduct scans in a safe, private space, and staff who are adequately trained and supported to use and interpret ultrasound images. National standards and training requirements for ultrasound use and associated care following ultrasound screening vary across countries, and influence routine ultrasound implementation.^11^ Many women, particularly those in limited-resource settings or who access antenatal care at primary health centres, lack access to routine antenatal ultrasound.^12,13^ In tackle this health care gap, the development of smaller, portable and handheld ultrasound devices is rapidly expanding, which could help improve women’s access to antenatal ultrasound in these settings.^14^

The application of artificial intelligence (AI) models and deep learning techniques have generated new approaches to interpreting medical imaging, including antenatal ultrasound.^15-17^ Algorithms that are trained on a database of high-quality, labelled ultrasound images (where the gold standard gestational age is known) can learn to estimate gestational age accurately, without the need for human expert measurement and interpretation of fetal biometry. Typically, such an AI model would ‘read’ images obtained through blind abdominal sweeps with a point-of-care ultrasound device. These sweeps can be performed by a person with minimal (rather than expert) training. Intelligent Ultrasound (IU, now part of GE Healthcare) is one of several companies that have developed an AI-based tool, called the the ScanNav FetalCheck system. This system estimates gestational age from ultrasound images obtained via the General Electric (GE) VScan^TM^ Air handheld ultrasound device.

### Rationale for a prospective cohort study on AI-based gestational age estimation in sub-Saharan Africa

Implementing an AI-based ultrasound will introduce new challenges for health services. While some women in Ghana, Kenya, and South Africa will have had a conventional ultrasound prior to being screened for eligibility to the study, approximately just under a half to three-quarteres of women will not, based on PEARLS site investigators’ estimates. Potential benefits of this AI-based ultrasound system for PEARLS include:

- AI-based ultrasound can be performed by a health worker with minimal (rather than specialist) training, so gestational age estimation can be scaled up more quickly and easily
- The gestational age estimate is generated by the software immediately and does not require expert review or interpretation.
- The IU ScanNav FetalCheck system uses GE’s handheld, point-of-care ultrasound device. The latter is commercially available, and has previously received regulatory approval in Ghana, Kenya, South Africa and many other countries.
- The hand-held, point of care ultrasound device allows easy transit between multiple sites, and the smaller size means it can be used in multi-purpose clinical areas.

The algorithm underpinning this functionality has been trained on a retrospective dataset of over 2 million antenatal ultrasound images from over 75,000 pregnant women, obtained from centres in Australia, India and the UK.^18^ It has been validated in silico using a separate retrospective validation dataset of over 700 patients, also from Australia, India and the UK. These studies showed the AI-based algorithm showed a mean absolute error of 1.7 days between 14-18 weeks gestational age and 2.8 days between 18-24 weeks, outperforming traditional fetal biometry.^18^

However, the IU ScanNav FetalCheck system has not been evaluated in a prospective study in real-world clinical settings for first or second trimester gestational age estimation or in African pregnancies and settings. That is, whether this system has similar (equivalent) accuracy to current standard-of-care using conventional ultrasound for pregnant women in PEARLS settings is currently unknown. If the system is shown to be at least equivalent to the current standard of care, the gestational age estimate it generates can be considered reliable for the purposes of informing FMF-based risk screening and aspirin implementation. In a mixed-methods formative research study, we will also identify challenges and the feasibility of implementing AI-based gestational age estimation using hand-held ultrasound (protocol published separately).^5^

## AIMS, HYPOTHESIS AND OBJECTIVES

### Primary aim

This study aims to assess the accuracy and precision of the IU ScanNav FetalCheck system (“IU system”) compared to conventional ultrasound for gestational age estimation, in pregnant women in Ghana, Kenya and South Africa.

### Primary objectives

1. Determine accuracy and precision of gestational age estimation by IU system in first trimester (11^+0^ to 13^+6^ weeks) as compared to gestational age estimation using CRL measurement by conventional ultrasound in first trimester (11^+0^ to 13^+6^ weeks).
2. Determine accuracy and precision of gestational age estimation by IU system in second trimester (14^+0^ to 27^+6^ weeks) as compared to gestational age estimation using CRL measurement by conventional ultrasound in first trimester (11^+0^ to 13^+6^ weeks).

### Secondary objectives

3. Determine accuracy and precision of gestational age estimation by IU system in second trimester (14^+0^ to 27^+6^ weeks) as compared to fetal biometry by conventional ultrasound in second trimester (14^+0^ to 27^+6^ weeks).
4. Determine the accuracy of identifying women at <20 weeks’ gestation by conventional ultrasound using second trimester fetal biometry, as compared to IU system in second trimester.

## METHODS

### Overview of study design

The prospective cohort study described in this protocol is one of several formative research activities conducted prior to the PEARLS trial (Figure 1). The additional research activities (protocols reported elsewhere) include mixed-methods research with health workers, women and health systems stakeholders to identify challenges and the feasibility of implementing pre-eclampsia risk screening and AI-ultrasound,^5^ and two prospective cohorts (one is Ghana, Kenya and South Africa,^4^ and a second in India) to evaluate the prognostic accuracy of a restricted variable-FMF risk screening tool.

This study uses a prospective cohort to evaluate accuracy of the IU ScanNav FetalCheck model at different gestational timepoints. The study will be implemented at selected study facilities in Ghana, Kenya and South Africa. Participating facilities are those with a trained, expert sonographer who will perform conventional ultrasound scans and fetal measurements (crown rump length [CRL], biparietal diameter [BPD], head circumference [HC], abdominal circumference [AC], femur length [FL] as required), in accordance with International Society of Ultrasound in Obstetrics & Gynecology (ISUOG) standards.^19,20^

### Study setting

The study will be conducted in Ghana, Kenya, and South Africa. In Ghana, University of Ghana Medical School investigators will collect data from one tertiary referral hospital, two district hospitals, and one primary health facility in the Greater Accra Region. In Kenya, University of Nairobi investigators will collect data from two tertiary/county referral hospitals and three secondary hospitals in Kiambu County, Kakamega County, and Kisumu City. In South Africa, University of Cape Town investigators will collect data from four midwife obstetric units in Cape Town’s Metro West area. The full list of facilities can be found in Appendix A.

### Training and Standardisation

All study sonographers will be accredited to ensure gold-standard measurements are standardised across study sites. At each participating facility, a local sonographer trained in conducting fetal ultrasound scans according to both national and ISUOG standards^19,20^ will be identified. Prior to the study commencing, the study sonographers will receive training on the PEARLS protocol and procedures from ultrasound advisors (expert clinician-trainers who are maternal-fetal medicine Consultants), as well as being provided with a detailed ultrasound training manual outlining exactly how each image should be acquired. Study sonographers will be accredited by these experts using standard quality assessment and criteria. All study scans (experimental and control) are performed using the GE VScan^TM^ Air ultrasound system (2-dimensional grayscale ultrasound with a curvilinear transabdominal probe).^21^ Following the training, each sonographer will submit 10 CRL images from scans conducted in the first trimester (<14 weeks) and 10 second trimester scans (containing the four biometric measurements: BPD, HC, AC, and FL) from scans conducted in the second trimester (14– 28 weeks). Each image will be reviewed by the Ultrasound Advisors and scored according to the INTERGROWTH 21st scoring criteria^22,23^ (Table 1). Individual scans will be deemed suitable for the study if for HC, BPD, AC and CRL the image has met at least four out of the six quality assessment criteria. For FL, a scan must meet at least two out of the four quality assessment criteria to be deemed suitable for study purposes. A study sonographer will be accredited only if over 80% of their submitted scan images meet the quality assessment standards. Where discrepancies in scoring arise, a third ultrasound advisor will review scans and differences in scoring resolved via discussion.

**Table 1:** Quality assessment criteria for scans obtained by study sonographers for accreditation by study Ultrasound Advisors. Quality assessment criteria are informed by INTERGROWTH-21^st^.^22,23^.

|  | Fetal biometric measurement |  |  |  |
| --- | --- | --- | --- | --- |
| Quality assessment | CRL | BPD/HC | AC | FL |
| **Criteria 1** | Mid-sagittal section | Symmetrical plane | Image shows the stomach bubble | Both ends of the bone clearly visible |
| **Criteria 2** | Fetus is in the horizontal orientation | Plane showing thalami | Image shows umbilical vein along 1/3 of the abdomen | >45 angle to the horizontal |
| **Criteria 3** | Fetus in neutral position | Cavum septum pellucidum 1/3 along midline echo | Kidneys not visible | Femoral plane occupying at least 50% of the total image size |
| **Criteria 4** | Fetus occupies at least 50% of the total image size | Cerebellum not visible | Multiple ribs not seen | Callipers placed correctly |
| **Criteria 5** | Crown and rump clearly seen and callipers are correctly placed | Fetal head occupies at least 50% of the total image size | Abdomen occupies at least 50% of the total image size |  |
| **Criteria 6** | Yolk sac is not | Callipers and dotted | Callipers / dotted |  |
|  | included | ellipse placed correctly | ellipse are placed correctly |  |

### Study population

The population eligible for this study are women presenting at a participating facility for an antenatal visit, between 11^+0^ to 13^+6^ weeks gestation. These women can be any age, parity, BMI and with a singleton or multiple gestation. Pregnant women in whom a major fetal abnormality is known or detected, or where a fetal heart rate cannot be detected, are not eligible.

### Recruitment

Women attending the participating facilities will be systematically approached by trained research staff and invited to take part in this study, in conjunction with other PEARLS formative research activities. Women with an estimated gestational age of <16 weeks based on best clinical estimate will be invited for screening. During the pre-screening process, an initial, routine conventional scan (gestational age, fetal heart rate) will be performed to assess eligibility. Those eligible for the study (gestational age 11^+0^-13^+6^, fetal heart rate present and no known congenital anomaly) will be invited to participate in the study and informed consent obtained prior to data collection. Women who are not eligible or do not consent to participate will receive the same clinical care they would otherwise receive. Study participants will also continue to receive usual clinical care. If an anomaly or absent fetal heart rate is detected during the study (or any other clinical issues are identified) the participant will be referred for appropriate clinical care.

### Gestational age estimation

#### Conventional ultrasound estimation (standard care)

At each study visit (first and second trimesters), study sonographers will use conventional ultrasound to assess gestational age as outlined in Box 1.

##### Box 1: Fetal biometry measurement practice for gestational age estimation

*Women in first trimester:*

- Abdominal ultrasound will be used to obtain three CRL measurements. The best measurement will be selected, and gestational age will be calculated using INTERGROWTH-21^st^ charts.^23^ The best CRL measurement by clinical judgement and associated image will be entered into REDCap.

*Women in second trimester:*

- At the second trimester ultrasound, BPD, HC, AC, and FL are each measured three times. The best measurement by clinical judgement of each (and associated image) will be entered into REDCap.
- Gestational age will be estimated using the INTERGROWTH-21 curves based on HC value (mm).^24^

#### Training on the IU FetalCheck device

Prior to the study commencing research midwives attended 1 day training, embedded within a 5-day in person training workshop on the PEARLS cohort and formative studies.^4,5^ Training consisted of an hour lecture on the theory of ultrasound and gestational age estimation, as well as the instructions for using the device, followed by hands-on training, using the device to conduct scans on pregnant women. Training was conducted by a member of the Intelligent Ultrasound team (Sacha Walton, Intelligent Ultrasound, GE Healthcare).

#### Intelligent Ultrasound gestational age estimation

A trained research midwife will complete six abdominal sweeps, as directed by video instruction using the IU FetalCheck device (Figure 2). The video directs the user on the correct orientation of the probe and maintaining full probe contact with the abdomen as well as guiding them on the sweeps. The required sweeps take an average of 1-2 minutes to obtain. The IU FetalCheck device has an inbuilt algorithm (version 3.0), which displays the estimated GA on the screen.

**Figure 2.**
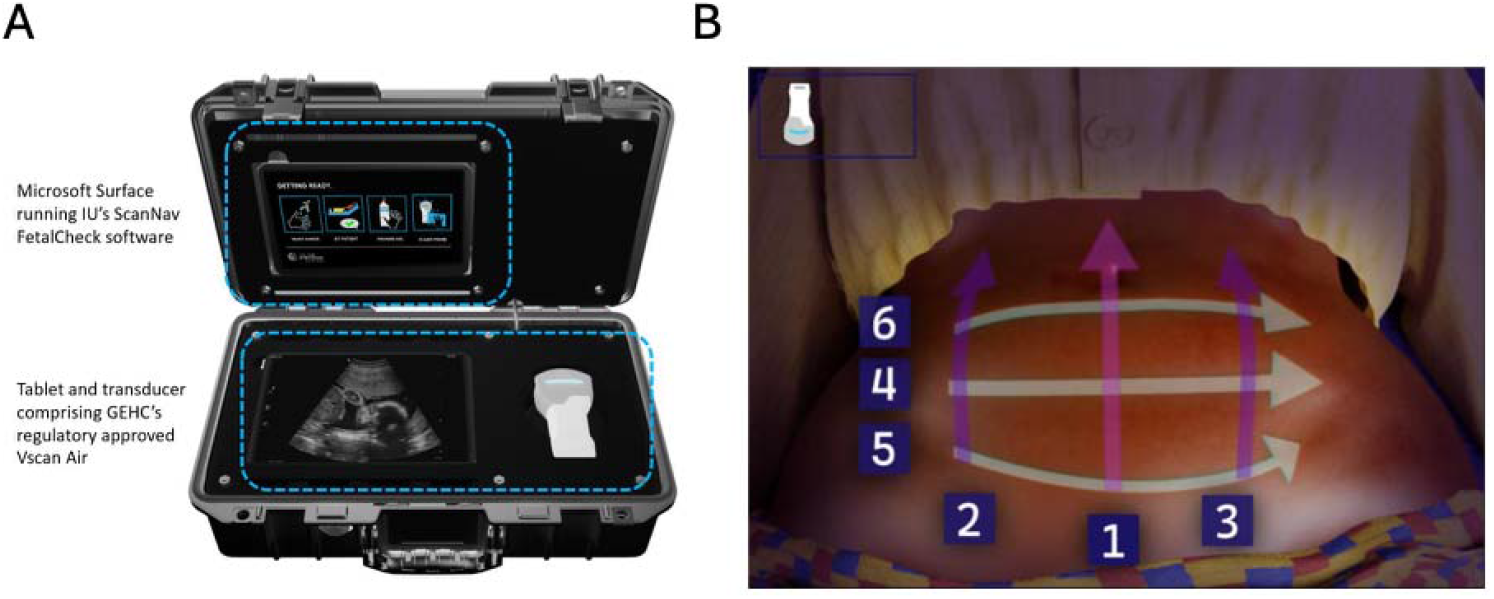
The IU ScanNav FetalCheck. (A) The upper portion of the device shows the IU ScanNav FetalCheck software is running on a Microsoft surface. On the lower portion contains the GE VScan^TM^ Air ultrasound system (2-dimensional grayscale ultrasound with a curvilinear transabdominal probe). (B) The visual instructions displayed by the IU ScanNav FetalCheck software, guiding the blind sweeps with the ultrasound probe.

### Data collection

REDCap, a Good Clinical Practice (GCP)-compliant, web-based data management platform will be used for this study. Screening for the study will be conducted via REDCap, and those eligible will be assigned a participant ID number. All study data included in the Case Report Form (CRF) as well as de-identified conventional ultrasound images, will be directly entered into REDCap using a secure tablet device.

### Participant flow and follow-up

The study comprises two visits for consenting participants (Visits #1 and #2; Figure 3). Following eligibility pre-screening (where the gestational age and viability of the pregnancy is confirmed), and informed consent is obtained, Visit #1 is conducted. Participants will have an abdominal ultrasound performed by a study sonographer using the GE VScan™ Air ultrasound system. The gestational age estimate from conventional ultrasound will be unblinded to the sonographer and participant (as per standard care). The CRL is measured 3 times and the best measurement by clinical judgement selected. Images from the best measurement will be saved, de-identified and uploaded to REDCap, along with the measurements obtained through the scan. Immediately after, a research midwife will perform a series of abdominal sweeps using the IU system, the estimated GA displayed on the IU system screen and the gestational age output from the IU system will be entered into REDCap by the midwife. The sonographer and research midwife will be blinded to each other’s gestational age results to minimise bias. Following the scans, baseline demographic data, blood pressure readings (using the CRADLE VSA device) and anthropometry will be collected from the participant, according to PEARLS procedures.^4^

**Figure 3.**
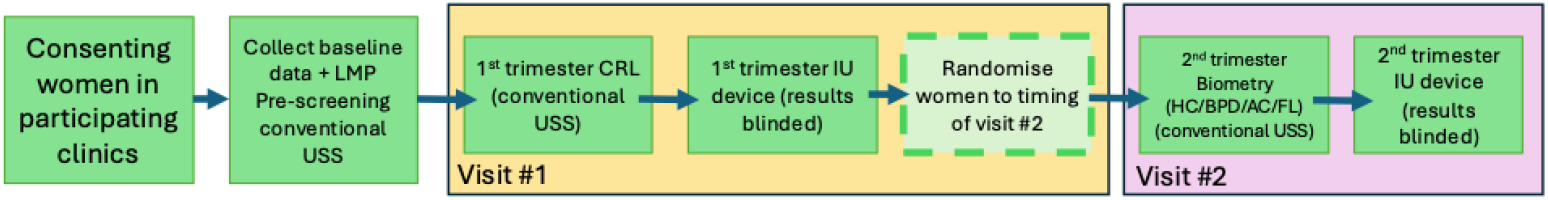
Flow of participants in the AI ultrasound validation study.

Women will be invited to return during the second trimester for Visit #2, at a randomly selected gestational age, between 14^+0^ and 27^+6^ weeks. The timing of Visit #2 is generated randomly in REDCap at the end of Visit #1. During Visit #2, the participant will again receive a conventional ultrasound, performed by a study sonographer. Again, the viability of the fetus will be confirmed via conventional ultrasound and any detected anomalies reported. Fetal biometry measurements will be obtained of the HC, BPD, AC and FL with the images and measurements uploaded directly to REDCap as well as a gestational age estimate according to the second trimester scan (by HC) Immediately after, a research midwife will perform a series of abdominal sweeps using the IU system, and the gestational age output will be entered into REDCap. A detailed description of the operational definitions of outcomes is included in Appendix B, Table 1. Where women cannot attend or miss their randomised appointment date (Visit #2, allocated to a 7-day period), women will be invited to attend at the closest available dates, as long as it is before 28 weeks.

### Statistical Methods and Analysis

#### Quality assessment

Throughout the study, the quality of images collected by sonographers on the conventional ultrasound will be assessed, to ensure high quality images are used for analysis of the gold-standard. For each sonographer, for the first 4 weeks of recruitment, all images will be assessed by the PEARLS ultrasound advisors, as per the criteria described in Table 1. For the remainder of the study 20% of the submitted images, selected randomly, will be assessed by the PEARLS ultrasound advisors. If the ultrasound advisors have concerns about the quality of scans from any of the sonographers during the first four weeks, that sonographers will continue to have 100% of images assessed throughout the study. Images that fail quality assessment will not be used for final analysis.

#### Sample size requirements

This study is powered at country level and is based on the 95% confidence intervals around the limits of agreement for each of the respective analyses are within +/- 10% of the values from previous studies. A total of 290 women are needed to achieve a minimum of 10% precision, meaning the sample size for Objectives #1 and #2 must each be at least 290 women. Each country will recruit until these targets are met. Assuming a 10% loss to follow-up between the first and second trimesters, each country will aim to recruit 323 women. We estimate the multi-country dataset will comprise 969 women.

#### Analysis

Baseline characteristics of study participants will be described using means and standard deviations for normally distributed variables, medians and 25^th^ and 75^th^ percentiles for non-normally distributed variables, and frequencies and percentages for binary and categorical variables.

Analysis for objectives 1, 2, and 3 will estimate (with 95% confidence intervals) the mean difference and 95% limits of agreement between the gestational ages estimates by IU and CRL (objective 1), ‘extrapolated CRL’ (objective 2), or fetal biometry (objective 3). Extrapolated CRL is the CRL-estimated gestational age plus the time difference (in days) between an individual’s first (visit 1) and second trimester (visit 2) measurement dates. The mean difference and 95% limits of agreement will be displayed with their 95% confidence intervals within a Bland-Altman style plot of the mean differences (y-axis) and average estimated gestational age (x-axis). In addition, fixed and proportional bias between gestational age estimates obtained using conventional ultrasound and the IU device will be evaluated using reduced major axis regression (RMAR).^25^ RMAR provides the slope and intercept of the regression line. A slope different from 1 indicates proportional bias, meaning that the magnitude of the difference between gestational age measured by conventional ultrasound and IU device varies with the magnitude of the conventional ultrasound measurements. If no proportional bias is present, an intercept different from 0 indicates fixed bias, meaning the difference between the two gestational age measurement methods remains consistent across the range of conventional ultrasound gestational age estimates.

Analysis of objective 4 will estimate (with 95% confidence intervals) the measures of diagnostic performance (specifically the detection rate and the false positive rate) of the IU gestational age estimation approach for classifying individuals as less than 20 weeks’ gestation, compared to conventional ultrasound fetal biometry, both measured at the second trimester, as binomial proportions. The complete data analysis plan is described in Appendix B.

#### Missing data

The proportion of missing data is likely to be low for objectives 1 and 3 given that both measurements are taken at the same time but may arise from technically difficulties such as electricity or equipment failure in the IU device. Objective 2 may have a small amount of missing data because of non-attendance for the second trimester follow-up (e.g. 10%), which we account for in the sample size calculation. Complete cases analysis will be used for all objectives. In the unlikely event of a high proportion of missing outcome data (e.g. over 10%) we will conduct a sensitivity analysis to evaluate whether there are meaningful patterns of missingness.

### Data quality assurance

Each study site will be responsible for data entry, as well as investigating and responding to data queries. The data management team will monitor range, consistency, and periodic reviews of distributions and identification of outliers. Regular feedback to sites will be used for quality assurance and to improve processes.

To ensure of ongoing adherence to the protocol and quality of images, a quality control system will be implemented. The intensity of the quality checks will vary across the life of the project depending on the quality of the scans received. For the first 25 scans from each sonographer, 100% of each conventional scan image will be reviewed by an ultrasound advisor, using the same criteria as the accreditation process (Table 1). If the study sonographer has provided ≥ 80% of images with suitable quality, for their next 25 scans only 50% will be reviewed. Subsequent image review will be determined by the quality of data received; this may vary by sonographer. For those sonographers where <80% of images are deemed suitable quality, feedback will be provided, and 100% image review will continue for the subsequent 25 scans. For sonographers where <50% of their image are deemed suitable quality, or if they have two scans where the images are unusable for gestational age estimation within a set of 25 scans, the sonographer will be immediately re-trained by both the local team and the Ultrasound Advisors. Data from each IU device will be periodically synchronised to a secure server, where the participant ID, timestamp, scan video and gestational age estimated by the IU system can be accessed by the data management team. This data is then matched to and cross-checked against the gestational age entered into REDCap to ensure accuracy.

### Ethical considerations

Research staff at study sites will be trained on GCP standards, including informed consent processes. An information sheet will be provided to eligible women, with non-technical and easy to understand language. These women will be given sufficient time to reflect on the information and will be given an opportunity to ask any questions.

If willing to participate, the woman will sign the consent form. This consent will be counter-signed by the research staff. Participants will be free to withdraw from the study at any stage without loss of benefits, and women will have the choice of including their data in analysis to the point of withdrawal, or exclusion of any data from analysis. If a woman cannot read or write, an impartial witness will be present during the informed consent process. The impartial witness will also sign and date the consent form, along with the individual who performed the informed consent discussion. Pregnant girls and adolescents who are eligible for this study and will complete an informed assent process. If an eligible participant is considered under the age of consent, her parent or other legally authorised adult will sign the informed consent, and the minor will sign the informed assent form. In South Africa, girls under the age of 16 years old were excluded, according to local ethical approvals. Participants will be provided with a study contact telephone number if they require further information or assistance. There will be no payment for participation, however some sites may reimburse participants for their travel costs.

## Discussion

The study described in this protocol forms part of the PEARLS formative research^4,5^ which aims to inform the design of the PEARLS randomised trial to identify the most effective and safest dose of low-dose aspirin for pre-eclampsia prevention. To implement this trial, accurate gestational age estimation is critical to 1) assess trial eligibility, 2) determine the primary outcome of preterm pre-eclampsia, and 3) accurately risk screen at enrolment. The prospective cohort described in this protocol will determine the accuracy of a novel AI-based system for gestational age estimation, that can be implemented with a commercially available hand-held ultrasound device used by non-expert operators. Furthermore, data collected will provide vital evidence for implementation of the WHO recommendation^11^ that all women should receive an ultrasound early in pregnancy, as it relates to gestational age estimation.

Currently, access to routine ultrasound for gestational age estimation remains inadequate in many LMICs.^12,13^AI-based ultrasound has the potential to overcome many of the known barriers to routine antenatal ultrasound.^13,26,27^ It removes the needs for specialised health care workforce to conduct ultrasounds and interpret the images. Growing evidence supports the efficacy, acceptability and feasibility of handheld ultrasound devices and AI-based estimation for gestational age in low-resource settings.^17,28^ Evidence from real-world clinical settings demonstrating the accuracy of AI-based ultrasound gestational age estimation would provide critical support for the wide-spread implementation of this technology, to improve equity of access and care of pregnant women globally. Validation of AI-based gestational age assessment could also open the door to further AI-based ultrasound assessments, such as fetal growth and well-being, placental and uterine assessments and prediction of pregnancy complications.

Accurate gestational age estimation is critical to the implementation of quality antenatal care, and numerous WHO recommended interventions to inform decisions on both maternal and neonatal care, including aspirin for the prevention of pre-eclampsia. Avalidated AI-based system to estimate gestational age will improve access to ultrasound-based gestational estimation, and risk screening for the prevention of pre-eclampsia.

## Data Availability

All data produced in the present work are contained in the manuscript

## List of abbreviations

FMF: Fetal Medicine Foundation
GCP: Good Clinical Practice
LMIC: Low- and Middle-Income Country
MAP: Mean Arterial Pressure
NCCEMD: National Committee for Confidential Enquiry into Maternal Deaths
NICE: National Institute for Health and Care Excellence
NPV: Negative Predictive Value
PAPP-A: Pregnancy Associated Plasma Protein-A
PEARLS: <u>P</u>reventing pre-eclampsia: <u>E</u>valuating <u>A</u>spi<u>R</u>in <u>L</u>ow-dose regimens following risk <u>S</u>creening
PLGF: Placental Growth Factor
PPH: Postpartum Haemorrhage
PPV: Postive Predictive Value
ROC: Receiver Operating Characteristic
TB: Tuberculosis
UTPI: Uterine artery Doppler Pulsatility Index
WHO: World Health Organisation

## Ethics approval and consent to participate

This study has received or sought ethics approval from the following entities: Australia: University of Melbourne, Office of Research Ethics and Integrity (Reference Number: 2024-28489-49438-3) and the Alfred Hospital Ethics Committee (Reference: Project 727/23); Ghana: Ghana Health Service Ethics Review Committee (GHS-ERC Number 002/01/24); Kenya: Kenyatta National Hospital, University of Nairobi ERC (Ref: KNH-ERC/01/MISC/20); South Africa: University of Cape Town, Faculty of Health Science, Human Research Ethics Committee (HREC Ref: 138/2024).

## Consent for publication

Not applicable

## Availability of data and materials

Not applicable

## Authors contributions

AS-D, ARAM, RCh, RCr, SJ, NJ, RKM, GG, DNK, LN, JAS, RH, ET, NM, AMG, AO, JPV and the PEARLS Collaborators made substantial contributions to the conception; AS-D, RCh, AO, AMcD, RCr, AMG and JPV design of the work; RKM, JAS and DNK design of statistical analysis, AS-D, AMcD, RCh, AMG, AO and JPV have drafted the work or substantively revised it. All authors reviewed and approved the submitted manuscript and have agreed both to be personally accountable for their own contributions.

## Acknowledgements

We would like to express our sincere gratitude to Manu Vatish (Gates Foundation) and the PEARLS Trial independent advisors (Cynthia Gyamfi-Bannerman, Eric Ohuma, Lloyd Tooke, Joyce N’gang’a, Koiwah Koi-Larbi Ofosuapea, Basky Thilaganathan, and Özge Tunçalp) for their support of this study. We thank Aris Papageorghiou (University of Oxford), as well as Nick Sleep and Sacha Walton (Intelligent Ultrasound, GE Healthcare) for device technical support and training. We also wish to acknowledge all members of the PEARLS Collaborators for their valuable contributions to the conduct of this study.

## PEARLS Collaborators

Alim Swarrey-Deen, Annie RA McDougall, Rosa Chemway, Rachel Craik, Skandarupan Jayaratnam, Naima Joseph, Robert K Mahar, Digsu N Koye, Long Nguyen, Julie A Simpson, George N Gwako, Ruth Hadebe, Edmund Tetteh Nartey, Nicole Minckas, A Metin Gülmezoglu, Joshua P Vogel, Ayesha Osman, Kwame Adu-Bonsaffoh, Evangel SA Amoah, Kwesi Aubin, Angeline Aywak, Kara Blackburn, Jennifer Blum, Meghan Bohren, James Chesang, Lester Chinery, Newlove Dadzie, Sue Fawcus, Alessandra Fleurent, Nchimunya Hapeela, Victoria Jepkosgei, Mushi Matjila, Jenipher E Okore, Samuel A Oppong, Alfred Osoti, Zahida P Qureshi, Belinda D Sackey, Jennifer Scott, Mark Sigei, Inge Smith, Marta Suarez-Fernandez, Saima Sultana, Ama A Tamatey, Nabeelah Toffar.

