## Supplemental appendix A for "Protocol for a prospective accuracy study on an artificial intelligence-based ultrasound system for gestational age estimation among pregnant women in Ghana, Kenya and South Africa"

### Appendix A - List of study sites

| Ghana | |
| --- | --- |
| Tertiary referral Hospitals | Greater Accra Regional Hospital |
| District Hospitals | Mamprobi Hospital |
|  | Ashaiman Municipal Hospital |
| Primary Health Facility | Tema Polyclinic |
| Kenya | |
| Tertiary Referral Hospitals | Kakamega County Teaching and Referral Hospital |
|  | Jaramogi Oginga Odinga Teaching and Referral Hospital |
| Secondary Level Hospitals | Kiambu Level 5 Hospital |
|  | Gatundu Level 5 Hospital |
|  | Ruiru sub-County Hospital |
|  | Malava sub-County Hospital |
| South Africa | |
| Midwife Obstetric Unit (MOU) | Mitchells Plain |
|  | Retreat |
|  | False Bay |
|  | Dunoon |
