## Supplemental appendix B for "Protocol for a prospective accuracy study on an artificial intelligence-based ultrasound system for gestational age estimation among pregnant women in Ghana, Kenya and South Africa"

Statistical Analysis Plan

| STUDY FULL TITLE | Formative research for the PEARLS Trial (Preventing pre-eclampsia: Evaluating AspiRin Low-dose regimens following risk Screening) - Prospective accuracy study on an artificial intelligence-based ultrasound system for gestational age estimation in Ghana, Kenya and South Africa |
| --- | --- |
| PROTOCOL VERSION | Version 3.0  Dated 5^th^ June 2024 |
| CHIEF INVESTIGATORS | Professor Joshua Vogel (Implementation Lead), Burnet Institute  Professor Metin Gülmezoglu (Sponsor), Concept Foundation |
| STATISTICIANS | Professor Julie Simpson, University of Melbourne  Dr Rob Mahar, University of Melbourne  Dr Digsu Koye, University of Melbourne |
| SAP AUTHORS | Professor Julie Simpson, University of Melbourne  Dr Rob Mahar, University of Melbourne  Dr Digsu Koye, University of Melbourne |
| SAP VERSION | 1.0 |
| SAP VERSION DATE | 19 December 2024 |

1 Table of Contents

### 2 Abbreviations and Definitions

| **Abbreviation** | **Definition** |
| --- | --- |
| AC | Abdominal circumference |
| AUC | Area under the receiver operating characteristic curve |
| BPD | Biparietal diameter |
| CRL | Crown rump length |
| DSMB | Data Safety Monitoring Boards |
| FL | Femur length |
| FMF | Fetal Medicine Foundation |
| GA | Gestational age |
| HC | Head circumference |
| IU | Intelligent Ultrasound |
| ICU | Intensive Care Unit |
| MISCH | Methods and Implementation Support for Clinical and Health research Hub |
| NPV | Negative predictive value |
| PEARLS | Preventing pre-eclampsia: Evaluating AspiRin Low-dose regimens following risk Screening |
| PPH | Postpartum haemorrhage |
| PPV | Positive predictive value |
| PO | Primary objective |
| ROC | receiver operating characteristic curve |
| SAP | Statistical Analysis Plan |
| SO | Secondary objective |
| TRIPOD | Transparent Reporting of a Multivariable Prediction Model for Individual Prognosis or Diagnosis |
| WHO | World Health Organisation |

### 3 Introduction

Pre-eclampsia is a multi-system disorder that develops during pregnancy due to abnormal placentation, dysregulation of angiogenesis, inflammation, oxidative stress, and maternal systemic vascular dysfunction. It is diagnosed through identification of new-onset hypertension in the presence of either proteinuria or new-onset maternal organ dysfunction at or after 20 weeks’ gestation. Pre-eclampsia is a leading cause of maternal and neonatal morbidity and mortality, globally it accounts for an estimated 14% of the 287,000 maternal deaths that occur each year.

Prophylaxis during pregnancy with daily low-dose aspirin is currently the standard treatment for women identified as high risk for pre-eclampsia. However, there is uncertainty surrounding the safest and most effective dosage of aspirin in low-, middle-, and high-income countries. In WHO’s 2021 recommendations on aspirin for pregnant women, the need for a randomised trial comparing a 150 mg dose to 75mg (current standard of care) was a high research priority. There is currently uncertainty as to whether the higher dose has greater benefit (greater prevention of pre-eclampsia) or may increase the risk of maternal bleeding.

This formative research project precedes, and will inform, the PEARLS (Preventing pre-eclampsia: Evaluating AspiRin Low-dose regimens following risk Screening) trial in Ghana, Kenya and South Africa. This formative phase aims to optimise gestational age (GA) estimation and identify potential barriers to trial implementation.

This statistical analysis plan (SAP) contains details of the planned statistical analyses to be completed to support the validation study of a new Artificial Intelligence (AI)-based tool for estimating gestational age, using point of care ultrasound, and forms part of the formative research of the PEARLS study.

### 4 Analysis Objectives and Endpoints

#### 4.1 Objective 1C - Accuracy and precision of the Intelligent Ultrasound

This study aims to assess the accuracy and precision of the Intelligent Ultrasound (IU) ScanNav FetalCheck system compared to conventional ultrasound for GA estimation, in pregnant women in Ghana, Kenya and South Africa.

**Primary objectives**

1C.PO1	Determine accuracy and precision of GA estimation by IU system in first trimester (<14 weeks) as compared to GA estimation using crown rump length (CRL) measurement by conventional ultrasound in first trimester (<14 weeks).

1C.PO2	Determine accuracy and precision of GA estimation by IU system in second trimester (14 to <28 weeks) as compared to GA estimation using CRL measurement by conventional ultrasound in first trimester (<14 weeks).

**Secondary objectives**

1C.SO1	Determine accuracy and precision of GA estimation by IU system in second trimester (14 to <28 weeks) as compared to fetal biometry by conventional ultrasound in second trimester (14 to <28 weeks).

1C.SO2	Determine whether there is a difference between women classified as <20 weeks’ gestation by conventional ultrasound using second trimester fetal biometry, as compared to IU system in second trimester.

1C.SO3	Explore the between- and within- rater variation in GA measured by GA estimation approach and by trimester, where possible.

#### 4.2 Endpoints

The endpoints for each of the objectives are listed below.

##### 4.2.1 Objective 1C - Accuracy and precision of the Intelligent Ultrasound

**Primary endpoints**

1C.PO1	GA estimated using the IU and conventional ultrasound during the first trimester (<14 weeks).

For first trimester IU, GA is estimated by the IU algorithm.

For the comparator (first trimester conventional ultrasound), GA is calculated by measuring CRL through abdominal ultrasound at <14 weeks. CRL is measured up to 3 times and the best measurement used. GA is then calculated using Robinson charts.

1C.PO2	GA estimated using the IU system during the second trimester (14 to <28 weeks) and extrapolated CRL GA by conventional ultrasound.

For second trimester IU, GA is estimated by the IU algorithm.

For the comparator, extrapolated CRL is the CRL-estimated GA from first trimester conventional ultrasound, plus the time difference (in days) between the individual’s first and second trimester measurement dates.

**Secondary endpoints**

1C.SO1-IC.SO2

GA estimated by the IU system in second trimester (14 to <28 weeks) and fetal biometry by conventional ultrasound in second trimester (14 to <28 weeks).

For second trimester IU, GA is estimated by the IU algorithm.

For the comparator (second trimester ultrasound scan), biparietal diameter (BPD), head circumference (HC), abdominal circumference (AC), and femur length (FL) will be measured. Each of these will be measured up to 3 times and the best measurement of each will be used. GA will be estimated using the Intergrowth-21 curves based on HC value (mm). Operational definitions for these endpoints are provided in Table 1 in the Appendix.

1C.SO3	GA estimated by the IU system and conventional ultrasound during the first and second trimesters.

### 5 Study Methods

#### 5.1 Study Design

We will conduct a prospective cohort study to confirm the accuracy and precision of the IU ScanNav FetalCheck system compared to conventional ultrasound for GA estimation, for pregnant women in the first and second trimesters. This study will recruit 969 women in Ghana, Kenya and South Africa who present for first antenatal visit prior to 14 weeks’ gestation. Women attending the participating antenatal clinics for their first antenatal visit will be systematically (consecutively) approached by trained research staff during clinic hours and invited to participate. For eligible women, the informed consent process will be undertaken before any data collection is performed.

The study comprises two visits for consenting participants. At Visit #1, participants will have baseline data collected and then undergo a dating scan performed by a sonologist using a standard ultrasound machine, with a GA determined using CRL. The GA estimate from conventional ultrasound will be unblinded to the sonologist and woman (as per standard care). Immediately after, a research midwife will perform a series of abdominal sweeps using the IU system, according to IU guidance.

Women will be invited to return during the second trimester for Visit #2, at a randomly selected GA (between 14 and 28 weeks). During Visit #2, they will again receive a conventional ultrasound (with GA calculated using fetal biometry) by an expert sonologist. Immediately after, a research midwife will perform a series of abdominal sweeps using the IU system.

### 6 Sample size

The sample size calculations are based on primary objectives 1 and 2 (1C.PO1 and 1C.PO2) specifically, that the 95% confidence intervals around the limits of agreement for each of the respective analyses are within +/- 10% of the values reported to the investigators by IU. A total of 290 samples are needed to achieve a minimum of 10% precision in objective 2, meaning the first objective sample size must be at least 290. Accounting for 10% loss to follow-up between the first and second trimesters, the sample size for objective 1 is 323. We will conduct this study in each of the three countries, hence the pooled (multi-country) dataset will comprise (3x323) 969 participants.

### 7 Analysis sets/Subgroups

Pregnant women who undergo GA estimation using both the IU ScanNav FetalCheck system (index test) and conventional ultrasound (reference test) during the first and second trimesters will form the analysis population. Women in whom a major fetal abnormality is known or detected will be excluded from the analysis.

Predefined sub-group analyses will be performed if sufficient data are collected, and will include by:

- country,
- maternal BMI (≥25, ≥30, ≥40 kg/m^2^)
- level of facility where scans are conducted (primary, secondary, tertiary).

### 8 Handling of Missing Values and Other Data Conventions

The proportion of missing data is likely to be low for objectives 1 and 3 (1C.PO1 and 1C.SO1) given that both measurements are taken at the same time. Objective 2 (1C.PO2) may have a small amount of missing data because of the second trimester follow-up (e.g. 10% at a maximum) and is accounted for in the sample size calculation for objective 1. Complete case analysis will be used for all objectives. In the unlikely event of a high proportion of missing outcome data (e.g. over 10%) we will conduct a sensitivity analysis to evaluate whether there are meaningful patterns of missingness to determine the best strategy for proceeding with the final analysis.

Participants who were enrolled to the study but did not meet the eligibility criteria or enrolled without an informed consent/assent will be excluded from the analysis.

### 9 Statistical Methodology

Baseline characteristics of all study participants included in each of the studies will be described using means and standard deviations for normally distributed variables, medians and 25^th^ and 75^th^ percentiles for non-normally distributed variables, and frequencies and percentages for binary and categorical variables.

For 1C.PO1, GA estimated by the IU will be considered as an index test and GA estimated using CRL measurement by conventional ultrasound in first trimester will be used as a reference test. To assess the accuracy and precision of GA estimation by IU system, the Bland-Altman analysis will be used to calculate the mean difference and 95% limit of agreement. The mean difference and 95% limits of agreement will be displayed with their 95% confidence intervals within a Bland-Altman style plot of the mean differences (y-axis) and average estimated GA (x-axis).

A similar approach to the primary analysis will be followed to address analysis objectives 1C.PO2 and 1C.SO1. GA estimated using the extrapolated CRL method will be used as a reference test for analysis objective 1C.PO2. Extrapolated CRL is the CRL-estimated GA plus the time difference (in days) between an individual’s first and second trimester measurement dates.

GA estimated using fetal biometry will be used as a reference test for analysis objective 1C.SO1.

Analysis of objective 1C.SO2 will estimate (with 95% confidence intervals), the measures of diagnostic performance (specifically the detection rate and the false positive rate) of the IU GA estimation approach for classifying individuals as less than 20 weeks GA, compared to conventional ultrasound fetal biometry, both measured at the second trimester, as binomial proportions.

Depending on availability of data from the training sessions of the intelligent ultrasound, analysis of exploratory objective 1C.SO3 will estimate the agreement in estimation of GA by within- and between-raters.

### 10 Sensitivity Analyses

No sensitivity analyses are planned.

### 11 Interim Analysis

A formal interim analysis will be performed following completion of enrolment of 100% of planned participants, and all available data on pregnancy and delivery outcomes. We estimate that ~67% of participants will have delivered by this time, though this is approximate if recruitment rates are slower or faster than expected. The results of the interim analyses will be shared with the Trial Steering Committee, and the Data Safety Monitoring Board (DSMB) prior to starting the PEARLS trial.

### 12 QC Plans

All data will be reviewed and cleaned before the start of any analysis. Data discrepancies will be recorded and resolved prior to undertaking the analysis. Any proposed changes to the data (e.g., due to implausible values identified for some variables) will be discussed with the team to reach consensus. All changes will be documented with rationale provided.

### 13 Programming Plans and allocation of who will complete the statistical analyses

Analyses will be undertaken using Stata/R. All analyses outlined in this statistical analysis plan will be undertaken by the University of Melbourne Methods and Implementation Support for Clinical and Health (MISCH) research Hub biostatistical team.

### 15 Appendix

**Table 1 – Outcome definitions for Objective 1C: the Intelligent Ultrasound (IU) device validation study**

| **Primary outcomes:** | **Operational definition and measurement** |
| --- | --- |
| 1^st^ trimester gestational age (conventional ultrasound) | 1^st^ trimester gestational age is calculated using the crown-rump length (CRL), measured by conventional ultrasound before 14 weeks, using the INTERGROWTH-21^st^ charts.  This is considered the ground-truth gestational age. |
| 1^st^ trimester gestational age (IU device) | Gestational age as estimated by the IU device before 14 weeks |
| Crown rump length (CRL) | The measurement (mm) from the endpoints of the crown to the rump, of a fetus orientated horizontally, in a neutral position (neither flexed or hyper-extended).  Best CRL measurement between 45 – 84 mm should be used for GA estimation. |
| 2^nd^ trimester gestational age (conventional ultrasound) | Gestational age is calculated using the head circumference (HC), measured by conventional ultrasound after 14 weeks, using the INTERGROWTH-21^st^ chart. |
| 2^nd^ trimester gestational age (IU device) | Gestational age as estimated by the IU device between 14 weeks 0 days and <28 weeks 0 days. |
| Biparietal diameter (BPD) | Measurement of BPD (mm) using the outer-to-outer placement of calipers (placement on the outer borders of the parietal bones at the widest part of the skull). |
| Head circumference (HC) | HC can either be measured using the ellipse approach or derived from BPD and occipital-frontal diameter (OFD) if the ultrasound device does not have ellipse function.  Ellipse approach: measured by placing the lone of the ellipse on the outer border of the skull (mm).  Derived approach: OFD measured using the outer-to-outer placement of calipers (placement on outer border of the occipital and frontal bones at the longest part of the skull).  HC (mm) = 1.62 x (BPD + OFD). |
| Abdominal circumference (AC) | The AC can either be measured using the ellipse approach or derived from linear measurements made perpendicular to each other, usually the anteroposterior abdominal diameter (APAD) and the transverse abdominal diameter (TAD)  Ellipse approach: measured (mm) by using the ellipse facility placed the line of the ellipse on the outer border of the abdomen.  Derived approach: APAD is measured with the calipers placed on the outer borders of the body outline, from the posterior aspect (skin covering the spine) to the anterior abdominal wall.  TAD is measured with the calipers placed on the outer borders of the body outline, across the abdomen at the widest point.  AC (mm) = 1.57 × (APAD + TAD). |
| Femur length (FL) | Measured (mm) by placing the the intersection of the calipers on the outer borders of the edges of the femoral bone (outer to outer). |

Note: all fetal biometry measurements are defined as per the recommended measurement techniques from ISUOG and INTERGROWTH.
